# “Double Machine Learning for Causal Inference in High-Dimensional Electronic Health Records”

**DOI:** 10.1101/2025.07.21.25331944

**Authors:** Mike Du, Yuchen Guo, Xintong Li, Marti Catala, Daniel Pareto-Alhambra

**Affiliations:** Pharmaco- and Device Epidemiology Group, Health Data Sciences, Botnar Research Centre, NDORMS, University of Oxford, United Kingdom; Department of Medical Informatics, Erasmus University Medical Centre, Rotterdam, The Netherlands

**Author notes:** **Corresponding author:** Name: Mike Du. **Funding:** This manuscript is funded by the Oxford NIHR Biomedical Research Centre.

**Keywords:** Double machine learning, Causal Inference, Epidemiology, High Dimensional Data, Machine Learning, Plasmode Simulations

## Abstract

**Background:** Estimating causal effects in observational health data is challenging due to confounding by indication. Traditional approaches such as inverse probability of treatment weighting (IPTW) rely on correct model specification, which is difficult in high-dimensional settings. We implemented an offset-based double machine learning (Offset-DML) practical framework for estimating binary treatment effects on the log-odds scale using logistic regression.

**Methods:** We have conducted a plasmode simulation study based on real-world clinical data, varying sample sizes (5,000, 10,000, 20,000) and outcome prevalence (5%, 10%, 20%) with 200 repetitions. We compared the performance of IPTW, stabilised IPTW, offset-DML (with and without cross-fitting), and high-dimensional DML (HD-DML). We measured and compared the performance of the different models with the following metrics: absolute bias, empirical standard error, and root mean square error relative to the true average causal effect.

**Results:** Across most scenarios, DML-based approaches outperformed IPTW methods in terms of bias and empirical standard error, particularly in larger sample sizes. Offset-DML showed comparable performance to HD-DML while avoiding convergence issues observed with HD-DML in sparse data settings. All DML methods had overlapping confidence intervals in most scenarios.

**Conclusion:** Offset-DML is a practical and robust alternative for causal inference in high-dimensional health data. Future work should investigate extensions to other outcomes and diagnostics to assess confounding control.

**Key messages:**

- Double machine learning based methods consistently outperform IPTW regarding bias and empirical standard error, particularly in large sample sizes and sparse-data scenarios.
- Offset Double machine learning is a practical and robust binary causal effect estimation method in high-dimensional settings.
- Unlike high-dimensional Double machine learning, the offset-based Double machine learning approach demonstrated consistent convergence across all scenarios, including those with low outcome prevalence and small sample sizes.

## Introduction

Electronic Health Records (EHR) data have become an important resource for conducting observational studies in epidemiology, providing routinely collected information on patient demographics, treatments, and outcomes (1). Unlike randomised controlled trials, which are often costly and time-consuming, analyses using EHR can be conducted more efficiently in real-world clinical settings. However, the observational nature of EHR data introduces several challenges, particularly when estimating causal effects in treatment comparative and safety studies due to confounding (2, 3).

One major challenge in controlling for confounding with EHR data is its high dimensionality. These datasets often include hundreds or thousands of covariates, covering patient characteristics, clinical measurements, and comorbidities (4, 5). While the large amount of data has the potential to account for all important confounders, it also creates issues such as overfitting and multicollinearity in traditional statistical methods for causal inference (6). Additionally, low sample sizes relative to the number of covariates are common, particularly in studies targeting rare conditions or treatments, further complicating causal effect estimation (7, 8).

Propensity score (PS)-based approaches, which are one of the most popular method in this field, face limitations in these high-dimensional settings. As these methods often rely on correctly specifying confounders through expert knowledge, a task that is often impractical in high-dimensional data settings (9, 10). Overfitting and multicollinearity of the propensity score model can further undermine the robustness of these approaches, leading to biased and unstable estimates of treatment effects (11).

Double Machine Learning (DML) has been proposed as a machine learning framework for causal inference in high-dimensional observational data, addressing many of these challenges with its flexible use of machine learning models (12-14). DML can accommodate complex, nonlinear relationships between covariates and outcomes by using machine learning algorithms to estimate nuisance parameters, which are model estimates related to treatment assignment and treatment outcome that are not of direct interest but are required for unbiased causal effect estimation. The method then employs orthogonalisation (15), to derive estimating equations that use these nuisance parameter estimates to directly partial out the confounding effects and produce unconfounded causal effect estimates (13). It also incorporates cross-fitting, a technique that partitions the data to separate model training from estimation (16, 17), minimising bias due to overfitting for causal effect estimations. However, its performance in high-dimensional EHR with binary outcomes has not been evaluated. Moreover, DML typically estimates causal effects on probability or risk difference scales, and obtaining interpretable odds ratios or log-odds treatment effects, which are often preferred in clinical research, often requires additional model transformations or post-estimation calculations.

Recent work by Liu et al. (18) has extended the DML framework to estimate causal effects on the odds scale directly. While this represents a methodological advance, their approach, like that of Chernozhukov et al. (13), requires solving semi-parametric estimating equations. This added complexity can make the method less accessible to applied researchers.

Here, we implemented an offset-based DML practical framework that uses a logistic regression model instead of semi-parametric estimating equations for causal effect estimation, with the estimates from the outcome and treatment assignment nuisance model. This design allows for directly estimating the causal effects on the log-odds scale without requiring post-estimation transformations. To evaluate the performance of the proposed practical framework, we conducted a plasmode simulation study with semi-synthetic data based on real-world EHR data, comparing its performance against more establish DML and propensity score-based methods. This study had two aims: first, to evaluate how DML methods perform in comparison to propensity score–based approaches commonly for observational studies with real-world EHR; and second, to assess the performance of our offset-based DML framework relative to more established DML approach for observational studies with binary outcomes and treatment assignment.

## Methods

### Simulation study design

We conducted a plasmode simulation study following White et al’s guideline (19) to assess the performance of the different approaches explained in this section under realistic clinical data conditions. Plasmode simulation is a semi-synthetic data generation approach that uses real-world datasets to create synthetic populations while preserving the complex relationships and correlation structures among covariates (20). This allows for evaluating different methods for causal effect estimations against known causal effects in settings that closely mimic observational studies with real-world EHR.

### Data generation process

The plasmode simulation was based on data from the Clinical Practice Research Datalink (CPRD) GOLD database (21) (see database description in supplementary material A), which contains a cohort of 149,082 patients newly diagnosed with psoriatic arthritis between 2000 and 2022. The patients in the cohort initiated one of four first-line treatments: methotrexate, sulfasalazine, leflunomide, or hydroxychloroquine (22). The observed treatment prevalences in the cohort were: methotrexate (41%), sulfasalazine (25%), leflunomide (8%), and hydroxychloroquine (26%). The CPRD data were mapped to the Observational Medical Outcomes Partnership (OMOP) Common Data Model (CDM) (23). We extracted 5,277 covariates from this dataset, including age at initiation of first-line treatment, sex, drug exposure history, and comorbidity history. Drug exposure and comorbidity covariates were defined as binary indicators, reflecting whether the individual had a record of the respective drug or condition at any time (−Inf, 0] before treatment initiation. Full details on cohort definitions are provided in Supplementary material B. A density plot of the prevalence of binary covariates in the dataset can bee see in Figure 1. The distribution is highly right skewed, indicating data sparsity as over 78% of covariates have a prevalence below 1%, and 95% fall below 5%.

**Figure 1:**
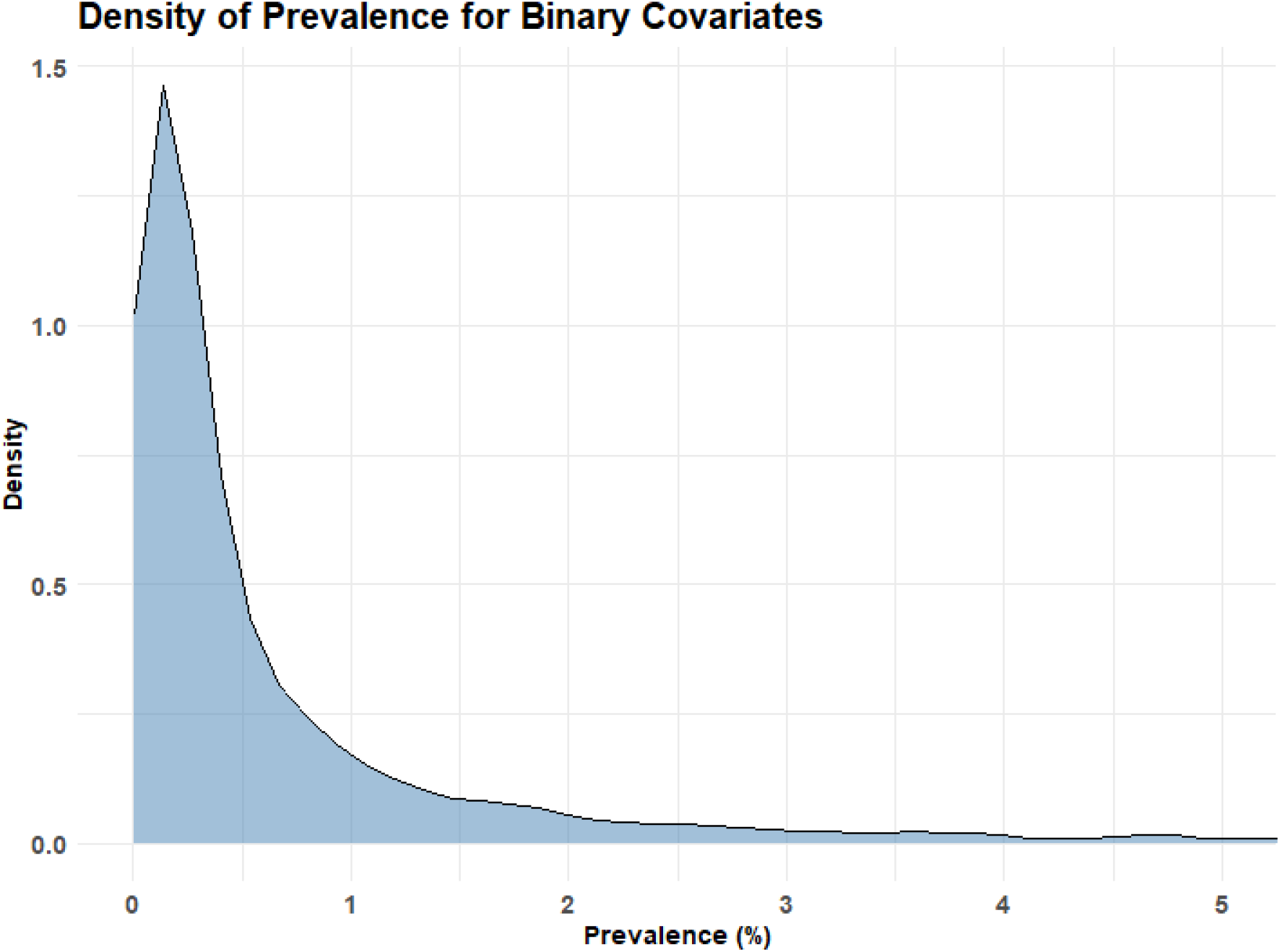
Prevalence of binary covariates of the CPRD dataset used for the plasmode simulation.

**Figure 1:**
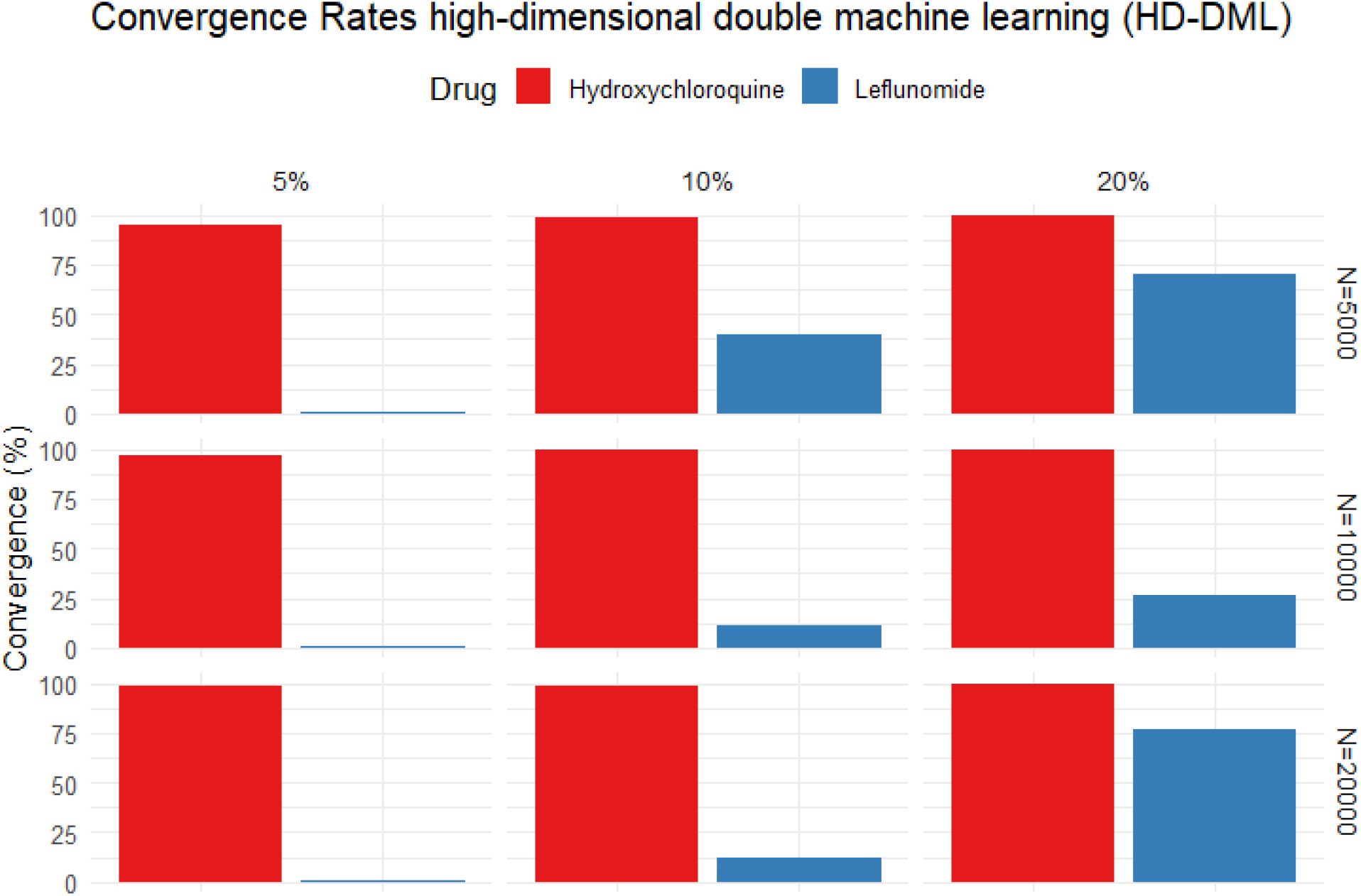
Convergence rates for high-dimensional double machine learning (HD-DML) for simulation scenarios varying by sample size, outcome prevalence, drug, exposure prevalence (prev), and true treatment effect. Other methods and results for Sulfalazine are excluded since all simulation iterations were converged.

Binary outcomes were generated using a linear logistic regression model with 500 covariates, age and sex, and an additional 498 random covariates from the full set of 5,277. This subset of 498 covariates was selected once at the beginning of the simulation and held constant across all repetitions within each simulation setting. This number was chosen to reflect the reality that, in clinical studies, only a small subset of covariates is typically expected to influence the outcome.

To assess method performance under varying outcome prevalence, we simulated outcomes at three different prevalence 5%, 10% and 20%, by adjusting the intercept of the outcome model. Prespecified log-odds true treatment effects were assigned as follows: sulfasalazine (–0.1), leflunomide (0.2), and hydroxychloroquine (0.1), with methotrexate as the reference.

For each outcome prevalence scenario, we examined three different total sample sizes 5,000, 10,000, and 20,000, to assess the impact of high dimensionality under different covariate to sample size ratios. For each of the nine resulting simulation settings, we generated 200 replicated datasets to evaluate the performance of the different methods compared in this study.

### Causal effect estimand

The causal effects estimand we estimated for this study is the average treatment effect (ATE) (24), which is the expected difference in outcomes between the treated and control groups. It can be expressed as follows:

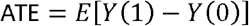

Where *Y*(1) and *Y* (0) denote the outcomes under treated and controlled, respectively.

### Offset-Based Double Machine Learning Framework for Log Odds Estimation

Let *Y* denote a binary outcome, *T* a binary treatment indicator, and *X* a high ⍰dimensional vector of covariates. We assume the partially⍰linear logistic model *logit P*(*Y* = 1|*T,X*) = *θT* +*r*_0_*(X)*, where *θ* is the causal log ⍰odds ratio of interest and the unknown function *r*_0_ (*X*) that captures covariate effects of *X*. The goal is to estimate *θ* through the double machine learning (DML) framework proposed by Chernozhukov et al (14), which follows the Neyman-orthogonal scores derived in Tan (25) and Liu et al (18).

We implemented an offset DML framework (offset-DML) to estimate *θ* which is the log-odds ratio of the binary treatment effect for the binary outcome *Y*. The framework contains three main steps with cross-fitting to reduce overfitting bias. The key idea here is to residualise both the treatment and outcome, and to use an offset term in the final model to isolate the causal effect of treatment on the log-odds scale with a logistic regression model.

#### Step 1: Residuals of Treatment Assignment Using Cross-Fitting

The treatment assignment is residualised in the first step to adjust for confounding by observed covariates. The dataset is randomly partitioned into equal-sized folds for cross-fitting, a procedure that reduces overfitting. On the training fold, we fit a logistic regression with LASSO regularisation of *T* on *X* restricted to observations with *Y* = 0. The fitted values 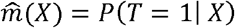 are predicted on the corresponding test fold and we then form the residualised treatment 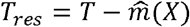 which removes the part of *T* that can be explained by observed covariates.

#### Step 2: Residuals of Treatment outcome Using Cross-Fitting

In the second step, the outcome is residualised following the same cross-fitting procedure. Using the same training data, we fit a logistic model of *Y* on (*T,X*), The fitted log⍰odds of the unknown 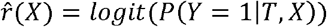 are estimated. This resulting log-odds values will serve as an offset term in the final regression model, aligning the residualised outcome with the log-odds scale used for treatment effect estimation.

#### Step 3: Logistic Regression with Offset Term

In the final step, on each held ⍰out fold we run, a logistic regression outcome model is fitted to estimate the treatment effect: 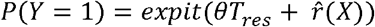. The residual treatment variable *T*_*res*_ from Step 1 is included as the independent variable, and the log-odds values 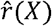 from Step 2 are included as an offset term, with the binary outcome from the dataset. Because the offset absorbs all variation in *Y* that can be explained by covariates, the single coefficient *θ* captures the remaining log-odds contribution of treatment. This procedure is algebraically equivalent to solving the Neyman-orthogonal score for the partially-linear logistic framework; the orthogonality property guarantees double robustness and, with cross-fitting, 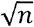 – consistent and asymptotically normal (18, 25).

This approach enables direct estimation of the average treatment effect on the log-odds scale, avoiding the need for post-estimation transformations. The resulting treatment effect estimate is statistically robust and directly interpretable as an odds ratio, which is commonly used in epidemiologic research (25). The fitting of nuisance models is flexible and can incorporate either traditional parametric models or machine learning algorithms, allowing for improved handling of complex, nonlinear relationships in high-dimensional data. This approach is consistent with the DML framework proposed by Chernozhukov et al.(13), which emphasises the importance of sample splitting and orthogonalisation to obtain unbiased estimates of causal effects, particularly in high-dimensional settings.

In the simulation study, both the outcome and treatment assignment nuisance models were estimated using a 5-fold cross-validated logistic Lasso regression (26).

### Benchmark Methods

To evaluate the performance for the Offset-DML, we compared it against three established methods for estimating average treatment effects: Inverse Probability of Treatment Weighting (IPTW), Stabilised IPTW, and the high-dimensional DML method proposed by Liu et al. (2021). Furthermore, we included a variant from our framework, Offset-DML without cross fitting (Offset-DML-no-cf) as a sensitivity to assess the impact on cross-fitting on the results for Offset-DML.

IPTW and Stabilised IPTW were included because they are among the most widely used methods in epidemiologic studies for confounding adjustment in observational data (27). Both were estimated using a propensity score model built with a 5-fold cross-validated logistic regression with Lasso regularisation. Average treatment effects were then obtained by fitting a weighted logistic regression of the binary outcome on the treatment assignment indicator, using weights defined by either the standard IPTW or the stabilised IPTW. Other weighting methods, such as overlap weights (28), have been shown to outperform IPTW approaches. However, they usually target a different estimand the average treatment effect in the overlap population, rather than ATE (29). Therefore, these methods were excluded to allow a fair comparison.

The high-dimensional DML (HD-DML) method proposed by Liu et al. was also included as a benchmark because, to our knowledge, it is the only existing DML approach specifically designed to estimate treatment effects on the log-odds scale in high-dimensional settings. This method uses Lasso regression models to estimate the treatment and outcome nuisance models, similar to the OFFSET-DML approach. Then, a preliminary treatment effect is estimated via a moment equation. In the final step, the outcome model is recalibrated using a regularised loss function that includes the initial estimate as an offset. A final treatment effect estimate is obtained using the updated equation incorporating the refined outcome model.

While conceptually related to our offset-DML framework, the Liu method differs in its use of a two-stage calibration procedure instead of cross-fitting to reduce overfitting bias in the treatment estimates. Full technical details for the Liu method can be found in Appendices 5 and 6 of Liu et al. (18).

## Performance Metrics

We assessed the performance of each method by computing the absolute bias (ABias) to measure the accuracy, empirical standard error (Emp-SE) and root mean square error (RMSE) to measure the precision of the estimated treatment effects relative to the known true effect. The formula for these metrics is included below:

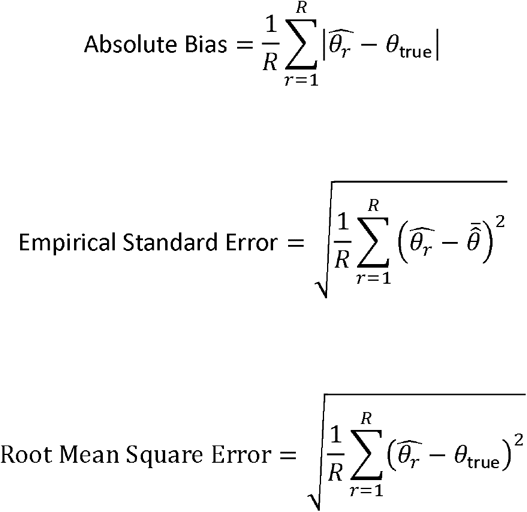

where 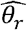 is the estimated treatment effect for simulation repetition r, R is the number of repetitions, *θ*_true_ is the true treatment effect and 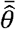 is the mean estimated treatment effect across simulations. The convergence rate of the methods was also reported.

### Implementation

The simulation study was implemented in R version 4.3.1, the Lasso regression for nuisance and propensity score model were implemented with glmnet package version 4.1-8 and the logistics regression outcome model were fitted with the stats package version 4.3.1. While the HD-DML method was fitted with the bespoke code provided from Liu et al.’s manuscript (18). The full analytical codes are provided on this github link.

https://github.com/oxford-pharmacoepi/DMLCausalInference

## Results

This section presents the results of five different methods evaluated in the simulation study: inverse probability of treatment weighting (IPTW), stabilised IPTW, offset double machine learning without cross-fitting (Offset-DML-no-cf), offset double machine learning with cross-fitting (Offset-DML), and high-dimensional double machine learning (HD-DML).

Convergence was achieved in 100% of iterations for all methods across all scenarios, except for HD-DML. As shown in Table 1, HD-DML exhibited poor or no convergence in multiple scenarios comparing leflunomide to methotrexate, with rates as low as 0.5% at the smallest sample size (n = 5,000) and only reaching 77% at the largest (n = 20,000). In contrast, convergence for HD-DML exceeded 95% in all scenarios involving hydroxychloroquine and sulfasalazine.

**Table 1:**
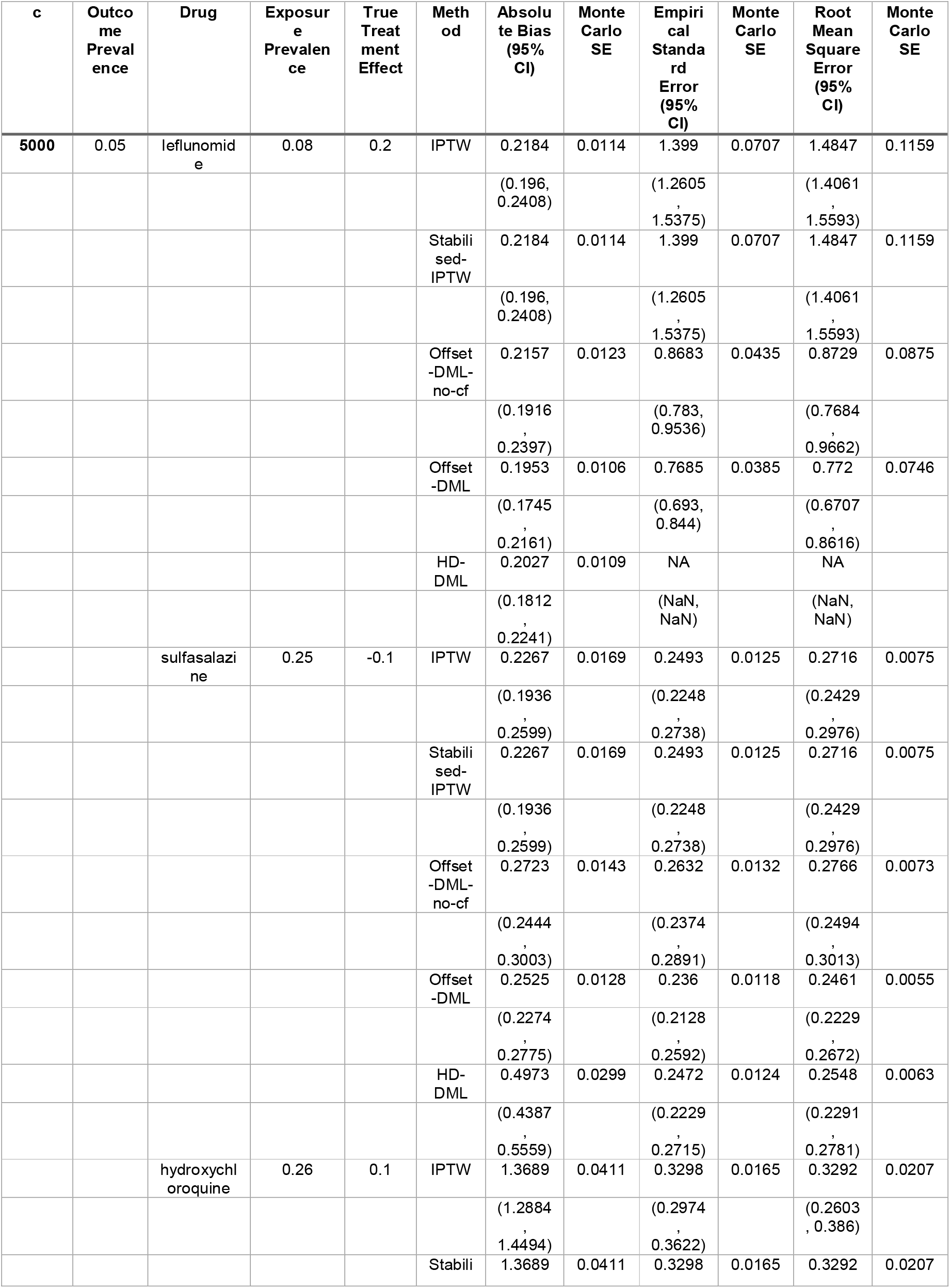
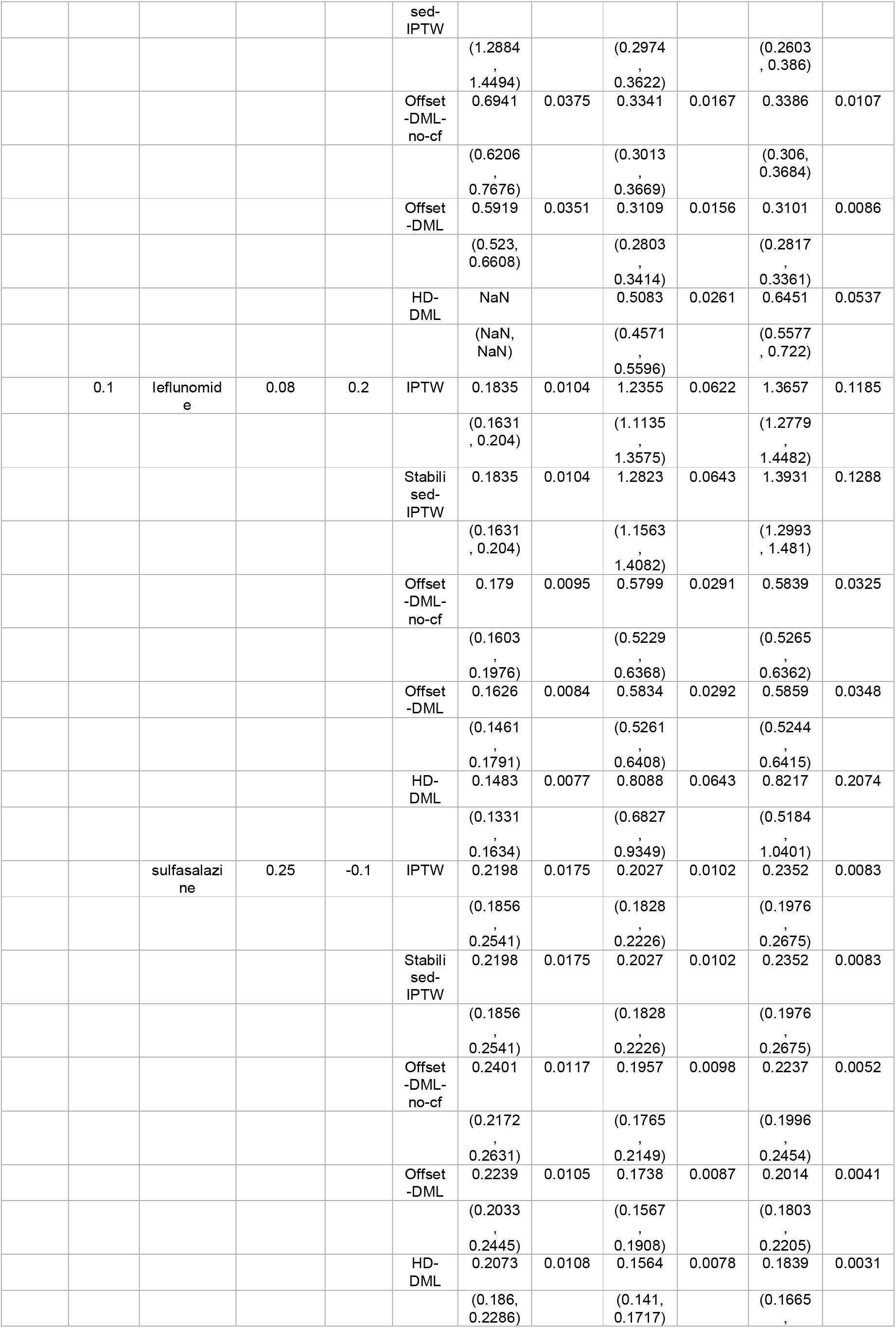
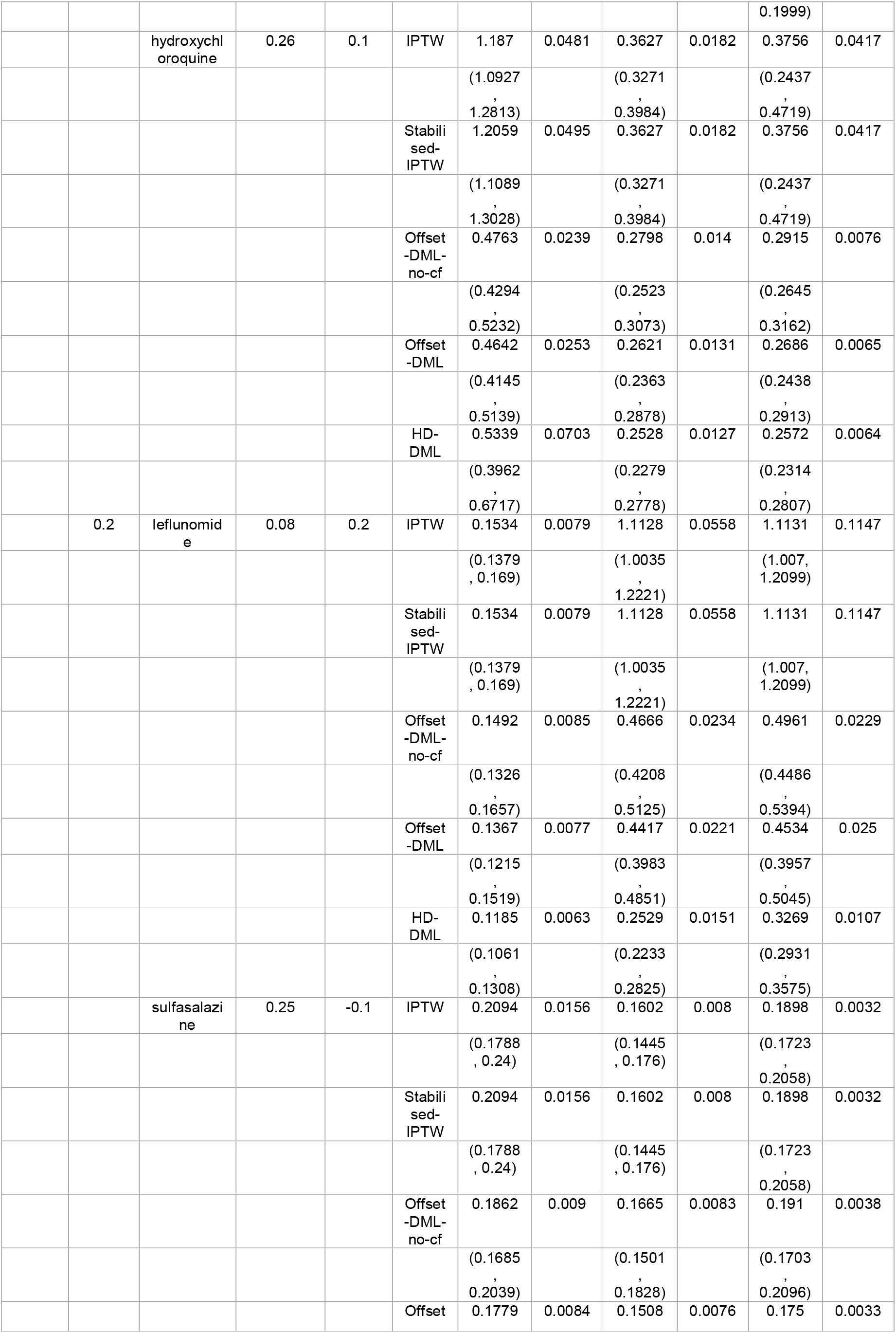
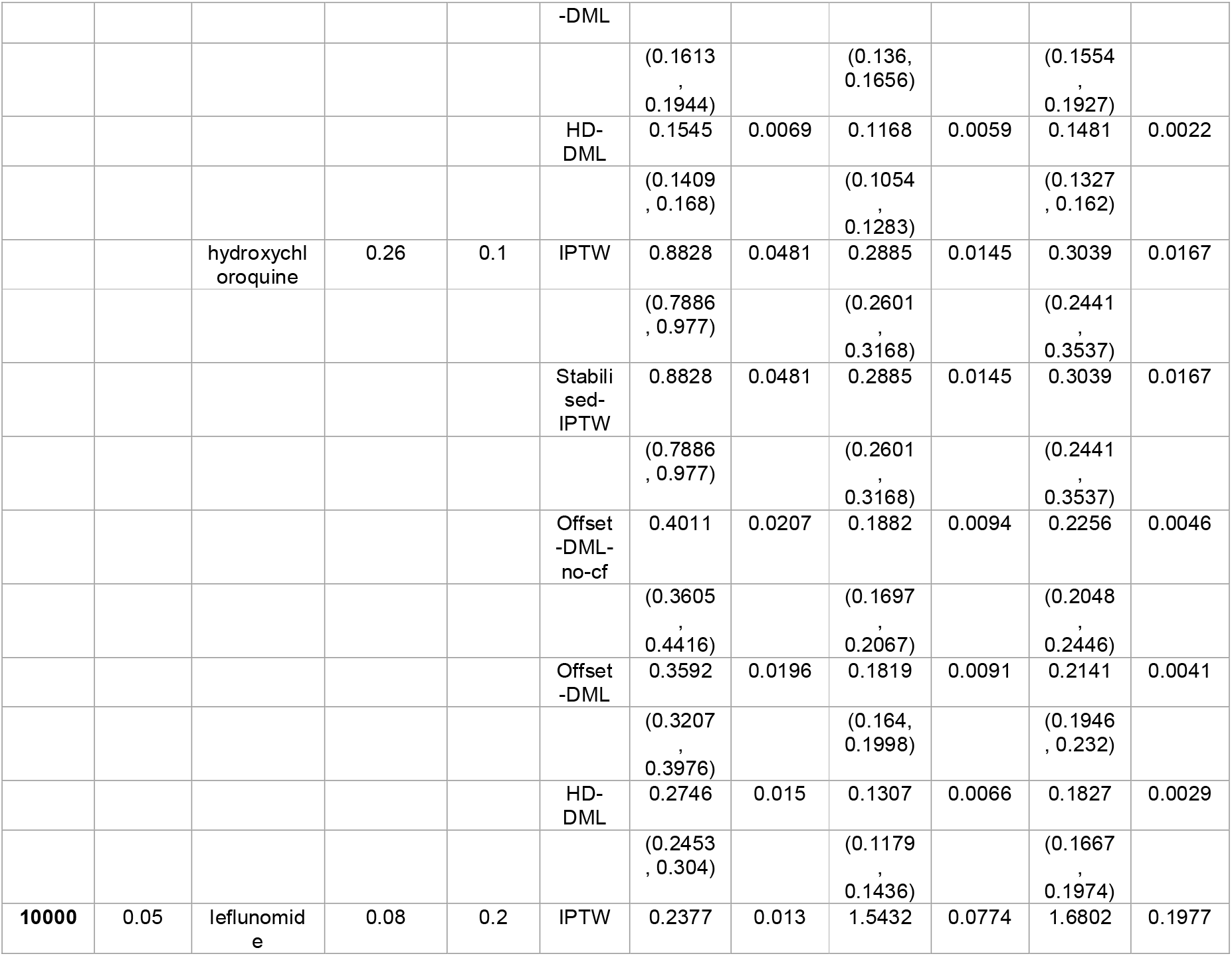
This table presents simulation results methods—IPTW, Stabilised-IPTW, Offset-DML (with and without cross-fitting), and HD-DML—under different conditions defined by sample size, outcome prevalence, drug, exposure prevalence, and true treatment effect. Performance metrics include absolute bias, empirical standard error, and root mean square error (RMSE), each with 95% Monte Carlo confidence intervals and Monte Carlo standard errors (MCSEs). Results are stratified by treatment scenario.

The ABias, Emp-SE, and Root-MSE are summarised in Figures 2–4 and table 1 shows the full result table. Across most scenarios, DML-based consistently yielded lower bias than IPTW-based methods. For instance, in the leflunomide scenario (outcome prevalence = 0.05; sample size = 10,000), the ABias was 1.40 (95% CI: 1.27 to 1.53) for IPTW, compared to 0.46 (95% CI: 0.41 to 0.51) for Offset-DML and 0.36 (95% CI: 0.33 to 0.40) for HD-DML. Similar patterns were observed across other drugs and sample sizes. Exceptions were observed at the smallest sample size (n = 5,000) for hydroxychloroquine and sulfasalazine, where the both methods performed similarly—for example, in the sulfasalazine scenario, the ABias was 0.22 (95% CI: 0.20 to 0.24) for IPTW and 0.20 (95% CI: 0.18 to 0.22) for HD-DML.

**Figure 2:**
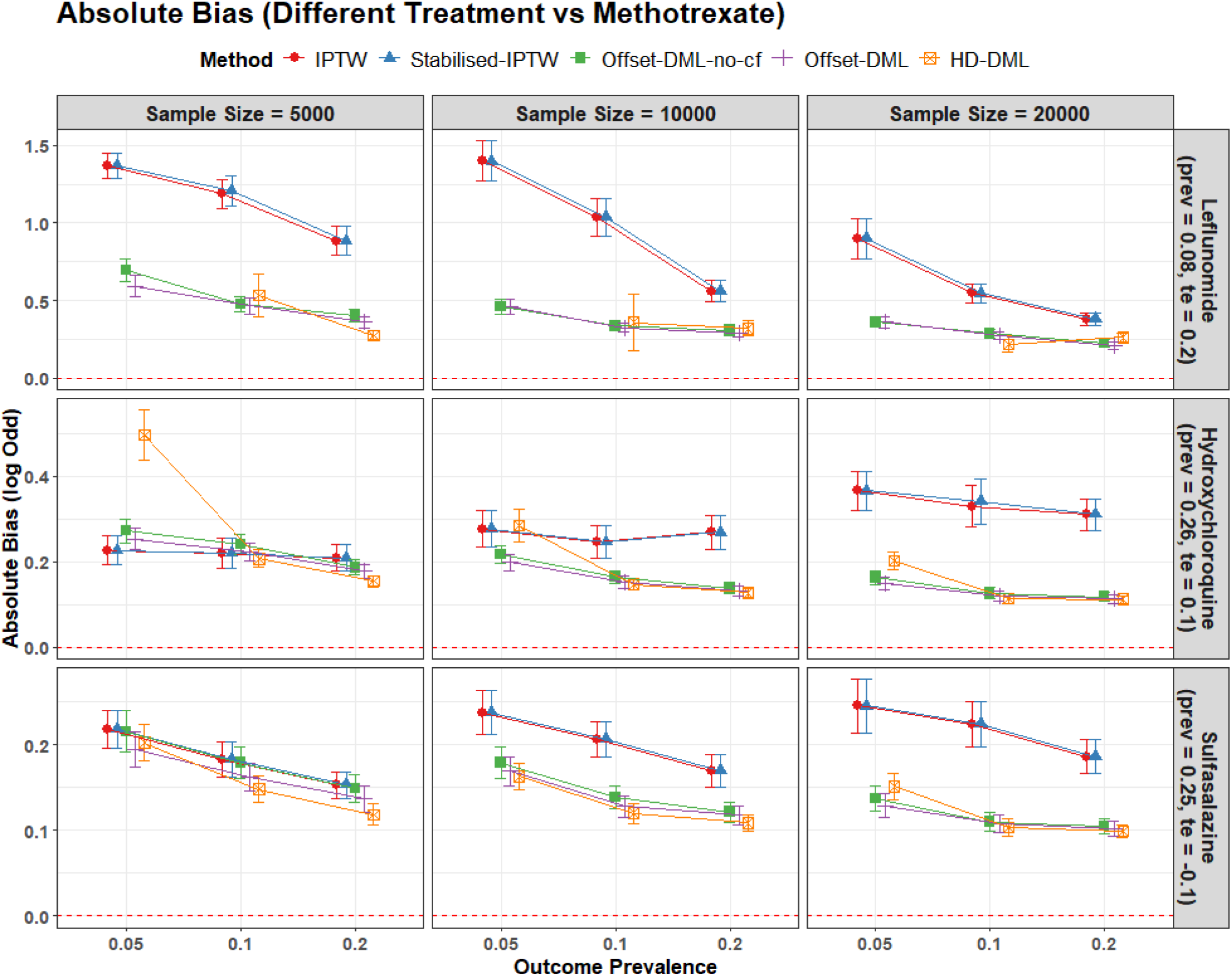
Average Absolute bias of five causal inference methods —inverse probability of treatment weighting (IPTW), Stabilised-IPTW, offset double machine learning without cross-fitting (Offset-DML-no-cf), offset double machine learning with cross-fitting (Offset-DML), and high-dimensional double machine learning (HD-DML) —for simulation scenarios varying by sample size, outcome prevalence, drug, exposure prevalence (prev), and true treatment effect (te). Values are reported as point estimates with 95% confidence intervals calculated with the Monte Carlo standard error.

**Figure 3:**
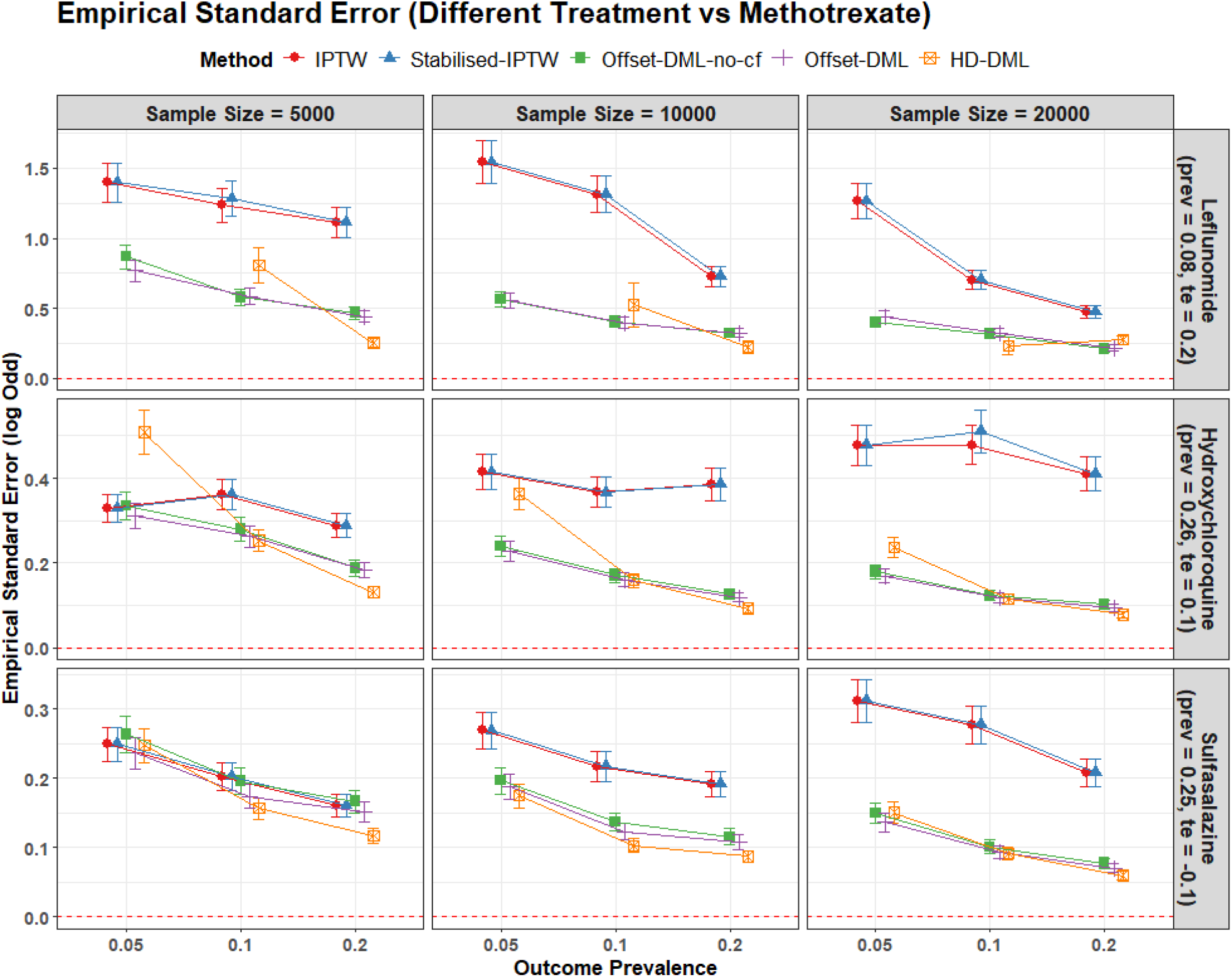
Average empirical standard error of five causal inference methods —inverse probability of treatment weighting (IPTW), Stabilised-IPTW, offset double machine learning without cross-fitting (Offset-DML-no-cf), offset double machine learning with cross-fitting (Offset-DML), and high-dimensional double machine learning (HD-DML) —for simulation scenarios varying by sample size, outcome prevalence, drug, exposure prevalence (prev), and true treatment effect (te). Values are reported as point estimates with 95% confidence intervals calculated with the Monte Carlo standard error.

**Figure 4:**
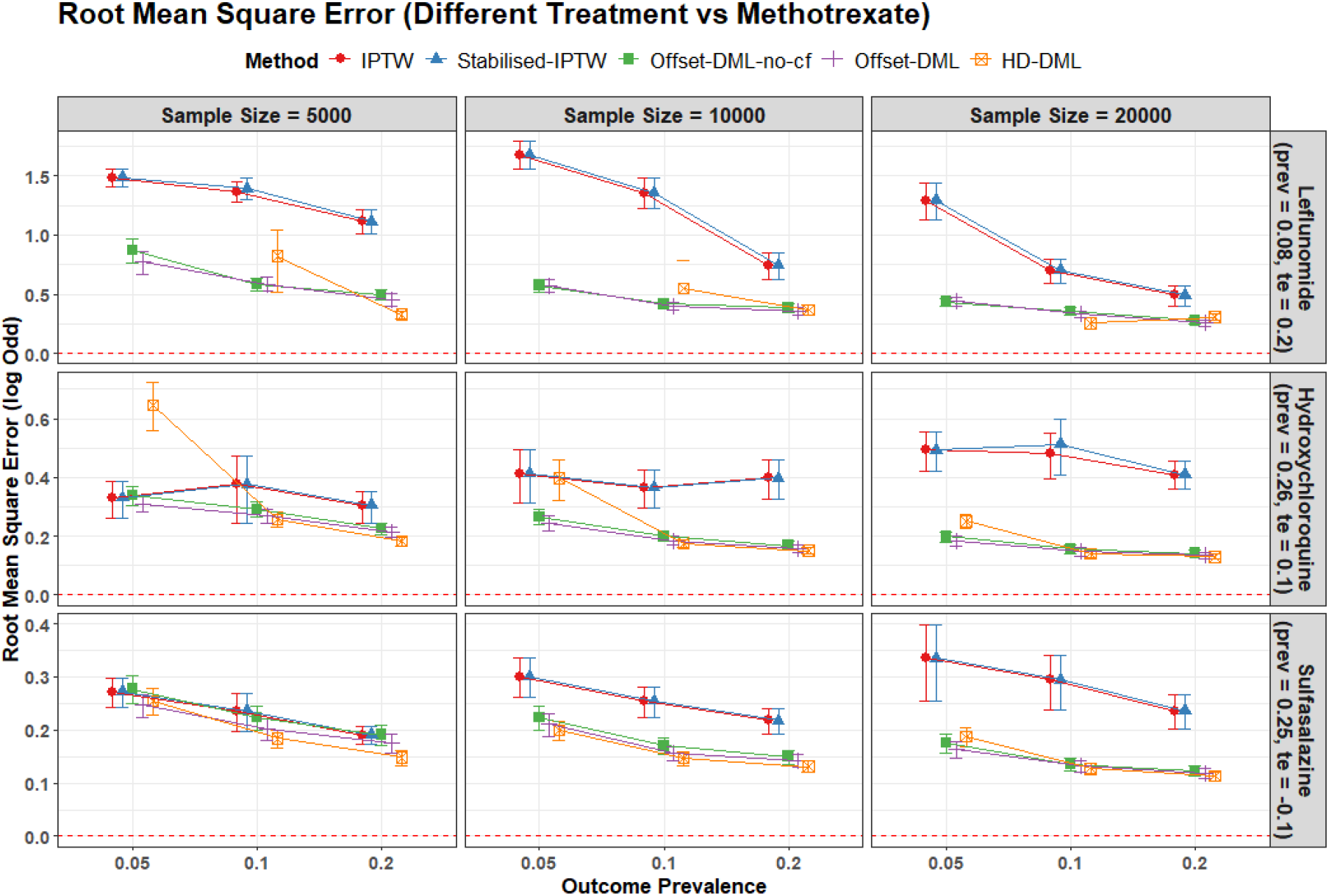
Average root square error of five causal inference methods —inverse probability of treatment weighting (IPTW), Stabilised-IPTW, offset double machine learning without cross-fitting (Offset-DML-no-cf), offset double machine learning with cross-fitting (Offset-DML), and high-dimensional double machine learning (HD-DML) —for simulation scenarios varying by sample size, outcome prevalence, drug, exposure prevalence (prev), and true treatment effect (te). Values are reported as point estimates with 95% confidence intervals calculated with the Monte Carlo standard error.

All the DML methods tend to give similar ABias with overlapping 95% CI in most scenarios, except in the low outcome prevalence (0.05), HD-DML tended to show higher bias than Offset-DML. For instance, in the hydroxychloroquine scenario (n = 5,000), the ABias was 0.50 (95% CI: 0.44 to 0.56) for HD-DML and 0.25 (95% CI: 0.23 to 0.28) for Offset-DML. Similarly, in the sulfasalazine setting, HD-DML had an ABias of 0.20 (95% CI: 0.18 to 0.22), while Offset-DML showed 0.16 (95% CI: 0.15 to 0.18).

Trends for Emp-SE and RMSE closely matched those of ABias. DML-based methods typically had lower empirical standard errors than IPTW-based methods. In the leflunomide example (n = 10,000; outcome prevalence = 0.05), Emp-SE was 1.54 (95% CI: 1.39 to 1.69) for IPTW, 0.56 (95% CI: 0.50 to 0.61) for Offset-DML, and 0.36 (95% CI: 0.33 to 0.40) for HD-DML. However, HD-DML often had the lowest empirical standard error in scenarios with high outcome prevalence (0.2). For instance, in the hydroxychloroquine scenario (n = 20,000; outcome prevalence = 0.2), HD-DML’s Emp-SE was 0.09 (95% CI: 0.08 to 0.10), compared to 0.12 (95% CI: 0.11 to 0.13) for Offset-DML and 0.39 (95% CI: 0.35 to 0.43) for IPTW.

## Discussion

In this study, we implemented an offset-based DML framework for estimating binary treatment effects on the log-odds scale. We evaluated its performance in a plasmode simulation study based on CPRD data that varied sample size, outcome prevalence, and exposure prevalence, reflecting a range of common high-dimensional clinical scenarios. The offset-based DML framework was compared to propensity score based IPTW, stabilised IPTW, and the high-dimensional DML method proposed by Liu et al. (18), which is designed explicitly for high-dimensional settings.

The simulation results showed that DML-based approaches consistently outperformed IPTW methods in terms of both bias and empirical standard error, with these differences particularly clear in scenarios with larger sample sizes (n = 10,000 and 20,000). Although both approaches used Lasso regression to accommodate high-dimensional covariates, IPTW depends on correctly specifying the propensity score model and including all relevant confounders. This requirement of the propensity score model can pose a challenge in high-dimensional settings, where knowledge-based confounder selection becomes impractical or infeasible (30). In contrast, DML-based methods do not require a prior identification of confounders. Instead, they achieve confounding adjustment through orthogonalisation that residual both the outcome and treatment assignment using separate nuisance models. This design helps to isolate the treatment effect and provides robustness against minor model misspecification (13, 18, 31).

Among the DML-based approaches, the proposed offset-DML method demonstrated comparable performance to the high-dimensional DML (HD-DML) approach introduced by Liu et al., both in terms of accuracy and precision. While HD-DML is theoretically appealing due to its tailored estimating equations for high-dimensional settings, the simpler offset-DML approach—relying on a standard logistic regression as the final model—performed just as well in our real-world clinical simulations. This highlights the practical advantage of offset-DML, particularly in sparse data scenarios commonly encountered in observational health data.

Furthermore, HD-DML experienced frequent convergence issues, particularly in the lowest exposure and outcome prevalence scenarios. This suggests that the estimating equations underlying HD-DML may be sensitive to numerical instability in sparse data and small sample settings (13). These challenges highlight a potential limitation when applying HD-DML in very high-dimensional and sparse settings, which are common in real-world epidemiologic observational studies (32, 33).

## Strength and Limitations

A major strength of this study is the use of a plasmode simulation based on real-world high-dimensional clinical data. This approach enabled us to evaluate causal inference methods under realistic and varied conditions, including differences in sample size, outcome prevalence, and exposure prevalence. The design reflects settings commonly encountered in electronic health records research and allows direct comparisons under controlled but representative scenarios.

However, the study has several limitations. First, while we employed Lasso regression for nuisance model estimation, we did not evaluate alternative machine learning algorithms that may further improve robustness in complex data settings. Second, even though we evaluated the methods in many scenarios, the findings may not generalise to settings outside those evaluated, for example, lower-dimensional datasets or data with less sparsity and different correlation structures.

## Recommendations for further research

From the limitations of the study, several topics remain for future research. These include exploring alternative machine learning algorithms for nuisance estimation, extending the method to other outcome types such as survival data, and developing framework for estimating model-based standard errors and compare its performance against other machine learning based causal inference methods such as Targeted Maximum Likelihood Estimation and Causal Forests (32). Additionally, research is needed to establish diagnostic tools that assess the extent to which DML approaches successfully reduce confounding bias in practice.

## Conclusion

In this study, we proposed and showed that the offset-DML gave comparable performance in terms of accuracy and precision to the high-dimensional DML approach by Liu et al (18), which was a more established DML approach, while avoiding the convergence issues that affected HD-DML in scenarios with low exposure prevalence and small sample sizes. It also consistently outperformed traditional IPTW approaches, particularly in larger sample sizes.

## Data Availability

All data produced in the present study are available upon reasonable request to the authors

## Supplementary Material

### A: Description of Data Sources Used for the Plasmode Simulation Study

The Clinical Practice Research Datalink (CPRD-GOLD) is a governmental, not-for-profit research service jointly funded by the National Institute for Health and Care Research (NIHR) and the Medicines and Healthcare products Regulatory Agency (MHRA), which is part of the UK Department of Health (https://cprd.com). CPRD-GOLD consists of computerized records of clinical and referral events in primary care, along with comprehensive demographic and prescription data for a representative sample of UK patients. The most recent data predominantly come from Scotland (52% of practices) and Wales (28% of practices).

The prescription records include detailed information such as the type of product, prescription date, strength, dosage, quantity, and route of administration. Data from contributing practices undergo quality checks at both the patient and practice levels during the initial processing stage. CPRD-GOLD provides data for over 20 million patients, including 3.2 million currently registered individuals. This study received approval through the Research Data Governance Process.

### B: Description of the cohort creation process for the plasmode simulation study

This section describe the participants included in the plasmode simulation study.

#### B.1. Description of the Cohort Creation Process for the Plasmode Simulation Study

The study cohort included adults newly diagnosed with psoriatic arthritis who initiated treatment with one of the following as there first-line therapies:

Methotrexate

Sulfasalazine

Leflunomide

Hydroxychloroquine

Participants were required to have at least one year of data available prior to the initiation of treatment to ensure sufficient baseline information.

#### B.2. Concept Codes Used to Identify Psoriatic Arthritis in the Database

The OMOP concept codes used to identify the psoriatic arthritis cohort in the database. Corresponding SNOMED codes can be accessed at https://athena.ohdsi.org/search-terms/start. Concept code = 19514005, 410482007, 33339001, 156370009, 239812005, 200956002, 10629311000119107.

#### B.3. Concept Codes Used to Identify Drug Cohorts in the Database

the OMOP concept codes used to define drug cohorts for the simulation study. Each cohort was created with all descendants of the specified drugs included. Corresponding SNOMED codes are available at Athena OHDSI https://athena.ohdsi.org/search-terms/start. Methotrexate (387381009), Sulfasalazine (387248006), Hydroxychloroquine (373540008), Leflunamide (386981009).

## References

1. Chute CG. Invited commentary: Observational research in the age of the electronic health record. Am J Epidemiol. 2014;179(6):759–61.

2. Austin PC. An Introduction to Propensity Score Methods for Reducing the Effects of Confounding in Observational Studies. Multivariate Behav Res. 2011;46(3):399–424.

3. Du M, Johnston S, Coplan PM, Strauss VY, Khalid S, Prieto-Alhambra D. Cardinality matching versus propensity score matching for addressing cluster-level residual confounding in implantable medical device and surgical epidemiology: a parametric and plasmode simulation study. BMC Med Res Methodol. 2024;24(1):289.

4. Schneeweiss S, Rassen JA, Glynn RJ, Avorn J, Mogun H, Brookhart MA. High-dimensional propensity score adjustment in studies of treatment effects using health care claims data. Epidemiology. 2009;20(4):512–22.

5. Stang PE, Ryan PB, Racoosin JA, Overhage JM, Hartzema AG, Reich C, et al. Advancing the science for active surveillance: rationale and design for the Observational Medical Outcomes Partnership. Ann Intern Med. 2010;153(9):600–6.

6. Schisterman EF, Perkins NJ, Mumford SL, Ahrens KA, Mitchell EM. Collinearity and Causal Diagrams: A Lesson on the Importance of Model Specification. Epidemiology. 2017;28(1):47–53.

7. Mitani AA, Haneuse S. Small Data Challenges of Studying Rare Diseases. JAMA Network Open. 2020;3(3):e201965–e.

8. Robertson SE, Leith A, Schmid CH, Dahabreh IJ. Assessing Heterogeneity of Treatment Effects in Observational Studies. Am J Epidemiol. 2021;190(6):1088–100.

9. Savitz DA, Wellenius GA, Savitz DA, Wellenius GA. 21Causal Diagrams for Epidemiologic Inference. Interpreting Epidemiologic Evidence: Connecting Research to Applications: Oxford University Press; 2016. p. 0.

10. Brookhart MA, Schneeweiss S, Rothman KJ, Glynn RJ, Avorn J, Stürmer T. Variable Selection for Propensity Score Models. American Journal of Epidemiology. 2006;163(12):1149–56.

11. Westreich D, Lessler J, Funk MJ. Propensity score estimation: neural networks, support vector machines, decision trees (CART), and meta-classifiers as alternatives to logistic regression. Journal of Clinical Epidemiology. 2010;63(8):826–33.

12. Bach P, Schacht O, Chernozhukov V, Klaassen S, Spindler M. Hyperparameter Tuning for Causal Inference with Double Machine Learning: A Simulation Study. In: Francesco L, Vanessa D, editors. Proceedings of the Third Conference on Causal Learning and Reasoning; Proceedings of Machine Learning Research: PMLR; 2024. p. 1065--117.

13. Chernozhukov V, Chetverikov D, Demirer M, Duflo E, Hansen C, Newey W, et al. Double/debiased machine learning for treatment and structural parameters. The Econometrics Journal. 2018;21(1):C1–C68.

14. Yuan J, Liu S. A double machine learning model for measuring the impact of the Made in China 2025 strategy on green economic growth. Scientific Reports. 2024;14(1):12026.

15. Neyman J, Scott EL. Consistent Estimates Based on Partially Consistent Observations. Econometrica. 1948;16(1):1–32.

16. Zivich PN, Breskin A. Machine Learning for Causal Inference: On the Use of Cross-fit Estimators. Epidemiology. 2021;32(3):393–401.

17. Newey WK, Robins JR. Cross-fitting and fast remainder rates for semiparametric estimation. arXiv preprint arXiv:180109138. 2018.

18. Liu M, Zhang YI, Zhou D. Double/debiased machine learning for logistic partially linear model. Econom J. 2021;24(3):559–88.

19. White IR, Pham TM, Quartagno M, Morris TP. How to check a simulation study. International Journal of Epidemiology. 2023;53(1).

20. Franklin JM, Schneeweiss S, Polinski JM, Rassen JA. Plasmode simulation for the evaluation of pharmacoepidemiologic methods in complex healthcare databases. Comput Stat Data Anal. 2014;72:219–26.

21. Delmestri A, Prieto-Alhambra D. CPRD GOLD and linked ONS mortality records: Reconciling guidelines. International Journal of Medical Informatics. 2020;136:104038.

22. Bhavsar SV, Movahedi M, Cesta A, Pope JE, Bombardier C. Retention of triple therapy with methotrexate, sulfasalazine, and hydroxychloroquine compared to combination methotrexate and leflunomide in rheumatoid arthritis. Joint Bone Spine. 2024;91(4):105732.

23. Hripcsak G, Duke JD, Shah NH, Reich CG, Huser V, Schuemie MJ, et al. Observational Health Data Sciences and Informatics (OHDSI): opportunities for observational researchers. MEDINFO 2015: eHealth-enabled Health: IOS Press; 2015. p. 574-8.

24. Rosenbaum PR, Rubin DB. The central role of the propensity score in observational studies for causal effects. Biometrika. 1983;70(1):41–55.

25. Tan Z. On doubly robust estimation for logistic partially linear models. Statistics & Probability Letters. 2019;155:108577.

26. Ranstam J, Cook JA. LASSO regression. British Journal of Surgery. 2018;105(10):1348-.

27. Xu S, Ross C, Raebel MA, Shetterly S, Blanchette C, Smith D. Use of stabilized inverse propensity scores as weights to directly estimate relative risk and its confidence intervals. Value Health. 2010;13(2):273–7.

28. Li F, Thomas LE, Li F. Addressing Extreme Propensity Scores via the Overlap Weights. American Journal of Epidemiology. 2018;188(1):250–7.

29. Austin PC. Differences in target estimands between different propensity score-based weights. Pharmacoepidemiol Drug Saf. 2023;32(10):1103–12.

30. VanderWeele TJ. Principles of confounder selection. Eur J Epidemiol. 2019;34(3):211–9.

31. Kabata D, and Shintani M. On propensity score misspecification in double/debiased machine learning for causal inference: ensemble and stratified approaches. Communications in Statistics - Simulation and Computation. 2025;54(5):1283–93.

32. Kim MK, Rouphael C, McMichael J, Welch N, Dasarathy S. Challenges in and Opportunities for Electronic Health Record-Based Data Analysis and Interpretation. Gut Liver. 2024;18(2):201–8.

33. Si Y, Du J, Li Z, Jiang X, Miller T, Wang F, et al. Deep representation learning of patient data from Electronic Health Records (EHR): A systematic review. Journal of Biomedical Informatics. 2021;115:103671.

